# *3D IntelliGenes:* AI/ML application using multi-omics data for biomarker discovery and disease prediction with multi-dimensional visualization

**DOI:** 10.1101/2025.03.25.25324634

**Authors:** Rishabh Narayanan, Elizabeth Peker, William DeGroat, Dinesh Mendhe, Saman Zeeshan, Zeeshan Ahmed

**Author notes:** **Corresponding author:** Zeeshan Ahmed, Rutgers Institute for Health, Health Care Policy and Aging Research, Rutgers University, 112 Paterson Street, New Brunswick, 08901, NJ, USA.

## Abstract

**Background:** The cutting-edge AI/ML techniques have proven effective at uncovering elucidative knowledge on disease-causing biomarkers and the biological underpinnings of a plethora of human diseases. However, the high-dimensional nature of multi-omics data presents numerous challenges in its effective presentation, annotation, and interpretation. Traditional 2D visualizations often fall short in capturing the intricate relationships between multi-omics features, hindering our ability to identify meaningful correlations.

**Methods:** In this study, we focused on addressing such challenges by developing an innovative solution to better visualize results produced by AI/ML approaches on integrated clinical and multi-omics data for novel biomarker discovery and predictive analysis. We present an advanced version of our earlier published software with intuitive and interactive visualizations of multi-omics data in multi-dimensions i.e., 3D *IntelliGenes*, which offers deeper insights, most importantly by capturing greater variability in the patient data by understanding both linear and non-linear structures, evaluating AI/ML model performance, and delineating the joint impact of biomarkers on the corresponding disease states.

**Results:** The overall functionality of 3D *IntelliGenes* is divided into two modules, data clustering and feature plotting. The data clustering module creates configurable 3D scatter plots to visualize the structure-preserving distribution of disease states, AI/ML classifier bias in the form of type I/II errors, and patient similarity through a robust density-driven clustering algorithm. Whereas the feature plotting module supports the joint analysis of pairs of multi-omics features to analyze the interdependence and discriminative power of co-expressed biomarkers.

**Conclusion:** We report evaluated performance of 3D *IntelliGenes* using diverse cohorts of patients with cardiovascular and other diseases.

## Background

The generation, management, integration, analysis, visualization, and sharing of multi-omics data, such as RNA-seq driven gene expression data, whole-genome sequencing (WGS) based variant data, and clinical and demographic-based patient data offer a holistic insight into the biological underpinnings of human disease, outperforming traditional single-omics analysis [1]. Furthermore, the application of artificial intelligence (AI) and machine learning (ML) techniques to integrative multi-omics analyses holds great potential to uncover disease-associated biomarkers and provide individualized treatment options to patients. However, the use of AI/ML techniques presents several challenges, including the need for computational expertise to handle large heterogenous datasets [2, 3]. Multi-omics data is inherently high dimensional in nature, making it difficult to visualize and interpret in a way that is inclusive of important structural details. Currently, two-dimensional (2D) plots currently constitute majority of the state-of-the-art visualizations for multi-omics data analysis e.g., Shapley Additive exPlanations (SHAP) scores, receiver operating characteristic (ROC) curves, confusion matrices, feature correlations using heatmaps, scatter and swarm plots and, and feature distribution plots, etc. [4]. 2D plots, however, often fail to capture high-dimensional structural details or discriminate between two patterns of equal importance, leading to analytic approaches that leave these excluded insights unexplored [5]. In this way, three-dimensional (3D) visualization allows for a deeper and more nuanced perspective of inter-feature relationships and can aid in understanding high-level trends and structures [6]. The development of interactive 3D visualization techniques can aid in revealing elucidative knowledge about human-disease etiology, and the lack of state-of-the-art tools in literature to take advantage of the benefits of 3D visualization for multi-omics leaves valuable room for improvement.

To support our ongoing exploration of the impact of integrative multi-omics analysis, we developed *IntelliGenes*, a novel AI/ML methodology combining a suite of classical statistical models with cutting-edge ML classifiers for disease prediction and biomarker discovery [7]. Our framework is based upon Findable, Accessible, Interoperable, and Reproducible principles for Research Software (FAIR4RS) approach and has been validated in peer-reviewed studies to outperform single algorithms in accurately predicting cardiovascular and other diseases [2], proving valuable for early diagnosis and personalized treatment. To provide more interactive visualization capabilities and to reduce barriers into complex, AI/ML-driven, integrative multi-omics analysis, we subsequently developed an accessible, user-friendly, and a cross-platform graphical user interface (GUI) of *IntelliGenes* to support researchers and clinicians with diverse backgrounds in appropriately and ethically applying AI/ML approaches to perform tailored analyses, discover novel biomarkers, and predict rare, common, and complex diseases with high accuracy [4, 7]. However, the current set of visualizations produced by *IntelliGenes* is restricted to solely 2D, which are impactful but suffer from the earlier mentioned limitations. Considering the advantages of 3D visualization and the current lack of 3D visualization tools for multi-omics data, in this article, we present 3D *IntelliGenes*, an extension of our methodology offering an interface for interactive 3D clustering and joint feature visualizations with implications in augmented/virtual reality (AR/VR). We strongly believe that 3D *IntelliGenes* can help better represent, understand, and interpret disease-specific structural details, inter-feature relationships, and AI/ML classifier performance for single-disease prediction. In this article, we present the methodology and user interface of 3D *IntelliGenes*. In addition, we validated the performance of 3D *IntelliGenes* on a diverse cohort of patients with cardiovascular diseases (CVDs) and reported the impact of multi-dimensional visualization on the interpretation of results produced using AI/ML approaches.

## Methods

3D *IntelliGenes* is composed of two modules: 1) AI/ML-Ready Data and Algorithms and 2) 3D Visualization, each containing various submodules. AI/ML-Ready Data and Algorithms interfaces support the user with data preparation, AI/ML configuration and analysis, and 2D Visualizations, while 3D Visualization interface enables the interactive visualization of results with submodules for Clustering and Feature Plots (Figure 1).

**Figure 1.**
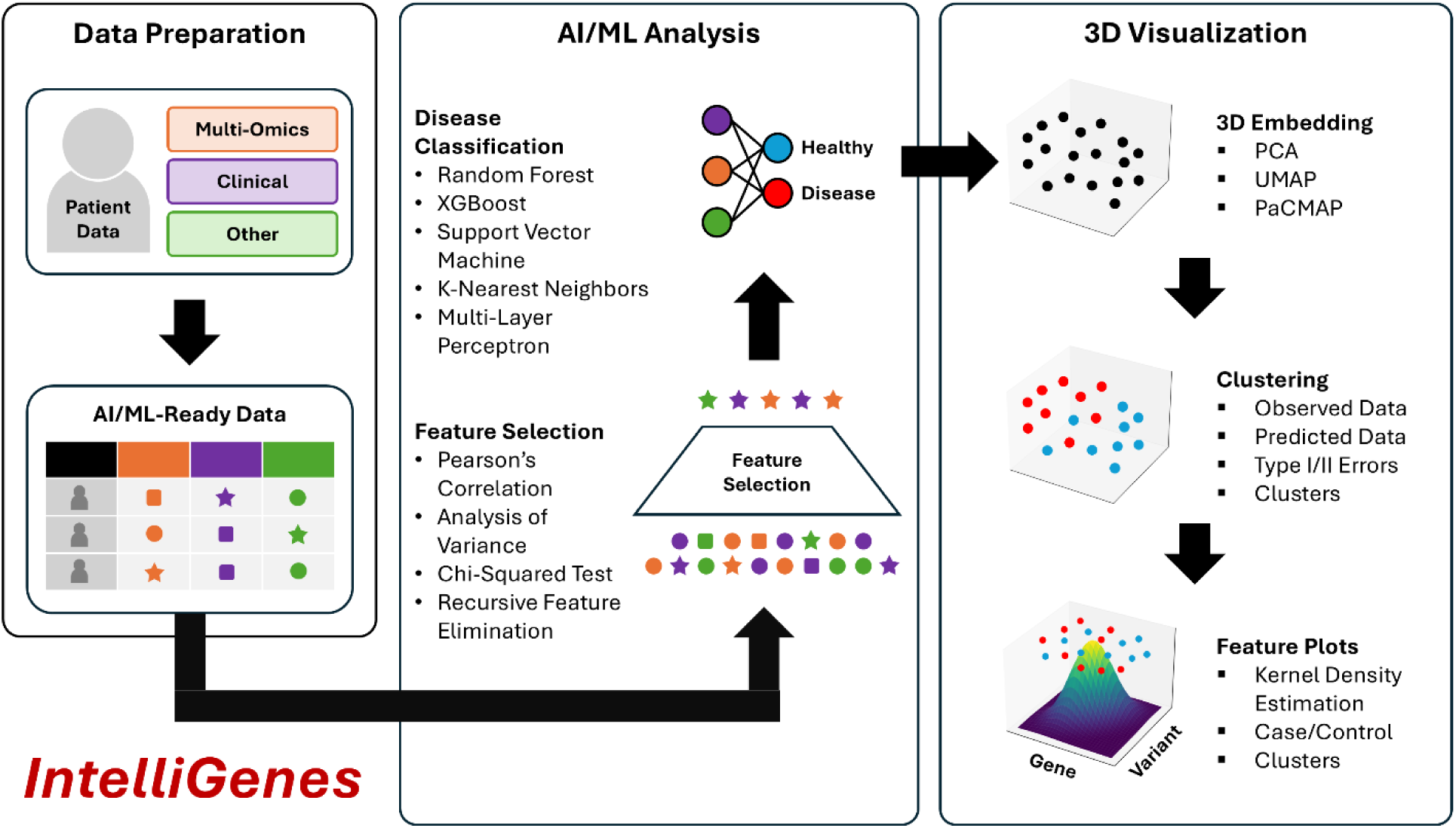
3D *IntelliGenes* workflow: 1) Data Preparation, 2) AI/ML Analysis, 3) 3D Visualization. Data Preparation enables the user in curating an AI/ML ready dataset. AI/ML Analysis offers robust and configurable feature selection and ML classification models. 3D Visualization provides three dimensional embeddings, clustering, and feature specific plots.

### AI/ML-Ready Data and Algorithms

Multi-omics data management, processing, and integration is inherently challenging due to its complex and heterogenous nature [8, 9]. Earlier, in addressing such challenges, we proposed and designed a new data format for extensibility, interpretability, and AI/ML readiness i.e., Clinically Integrated Genomics and Transcriptomics (CIGT). It supports structuring various genomic, transcriptomic, and clinical and demographic features, which can be then used for AI/ML-based predictive analysis.

*IntelliGenes* implements a dual-stage technique for biomarker discovery and AI/ML classification for single-disease prediction based on a collection of classical statistical techniques and ML classifiers. To identify significant features for predicting disease, *IntelliGenes* combines results from four separate selector models: Pearson’s correlation, chi-square (*χ*^2^) test, analysis of variance (ANOVA), and recursive feature elimination (RFE). Only features found significant in all tests are preserved for subsequent classification. *IntelliGenes* then uses these models to train a user-driven ensemble of five ML models: random forest (RF), support vector machine (SVM), Xtreme Gradient Boosting (XGBoost), k-nearest neighbors (KNN), and multi-layer perceptron (MLP), which are compiled downstream by a voting classifier i.e., Intelligent Gene (I-Gene) score. Through *IntelliGenes*, the user can configure and tune various hyperparameters to improve model accuracy. We published a study describing the advantages and disadvantages of different ML algorithms and their application to various diseases, which could be considered in the configuration of the software [10].

The overall graphical interface of 3D *IntelliGenes* adapts the original framework of the earlier published version of the *IntelliGenes GUI* [4]. This includes three submodules: 1) Data Manager, 2) AI/ML Analysis, and 3) Visualization. The Data Manager module supports the user in preparing and editing their CIGT-formatted AI/ML ready multi-omics dataset and in choosing an output directory to save all generated results. The AI/ML Analysis module enables the selection of statistical-based selection models and ML classifiers through an interactive and extensive configuration panel. It also supports the execution of AI/ML analysis and the monitoring of progress through a live console. Lastly, the Visualization module supports the user in filtering results generated during analysis and offers a convenient viewport to inspect results directly within the interface, including all the 2D visualization options (e.g. SHAP scores, ROC curves, confusion matrices, feature correlations using heatmaps, scatter and swarm plots and, and feature distribution plots) generated by *IntelliGenes*. Here, after completing AI/ML analysis, the user is also presented with an option to launch the interactive 3D Visualization module in a separate window.

In embodying FAIR4RS principles, we have open-sourced the code for our 3D *IntelliGenes* platform like earlier versions of the IntelliGenes [4, 7]. 3D *IntelliGenes* is developed in the Python programming language and relies on standardized, widespread, and state-of-the art libraries. To transform and manipulate the data for analysis and visualization, we use *Pandas* and *numpy*. The *scikit-learn*, *SciPy*, and *XGBoost* libraries are used for the various statistical and ML algorithms central to the *IntelliGenes* methodology. *SHAP* is used to generate interpretable feature importance scores for otherwise black-box ML methods. The diverse set of 2D and 3D visualizations are generated using *Matplotlib* and *seaborn*, and *Qt* with *PySide6* is used to render cross-platform GUI components. To package the executable across all major operating systems (i.e., MacOS and Windows), we used *PyInstaller*.

### 3D Visualization

Data visualization is a crucial aspect of exploratory data analysis and data interpretation. Visually understanding the distribution and inter-relationships of multi-omics data can provide deeper insights into the underlying biological pathways and structures observed in the data. The first and second released versions of *IntelliGenes* offer 2D visualizations due to the general simplicity and widespread accessibility of such plots [4, 7]. Each plot offers a specialized interpretation of the underlying dataset, enabling the user to understand metrics such as AI/ML model performance, data distribution, and inter-relationships. Furthermore, during the development of first two versions of *IntelliGenes*, we were more focused developing a customizable AI/ML data analysis approach. However, moving forward we are keen on improving the annotation, interpretation, and presentation of AI/ML results with better visualization techniques [4, 7].

2D visualizations of high-dimensional multi-omics data add great value to data interpretation and presentation, however, this approach is inherently limited due to its inability to explain finer structural nuance and inefficacy in discriminating between two equally important patterns [5]. Extending 2D into 3D can capture greater proportions of multidimensional variability in the original dataset and can better represent underlying structures. Although 3D visualizations have seen advances in fields like medical imaging (e.g., cancer detection) [11], protein folding [12], and chemical space visualization [13], it has yet to see similar advancements in the realm of multi-omics. In 3D *IntelliGenes*, we implement a robust 3D visualization workflow comprised of two important stages to explore the structure and inter-relationships of the data respectively: 1) Clustering and 2) Feature Plots (Figure 2). Each stage is contained within a separate tab in the 3D *IntelliGenes* application window, which may be launched directly from the Visualization module.

**Figure 2.**
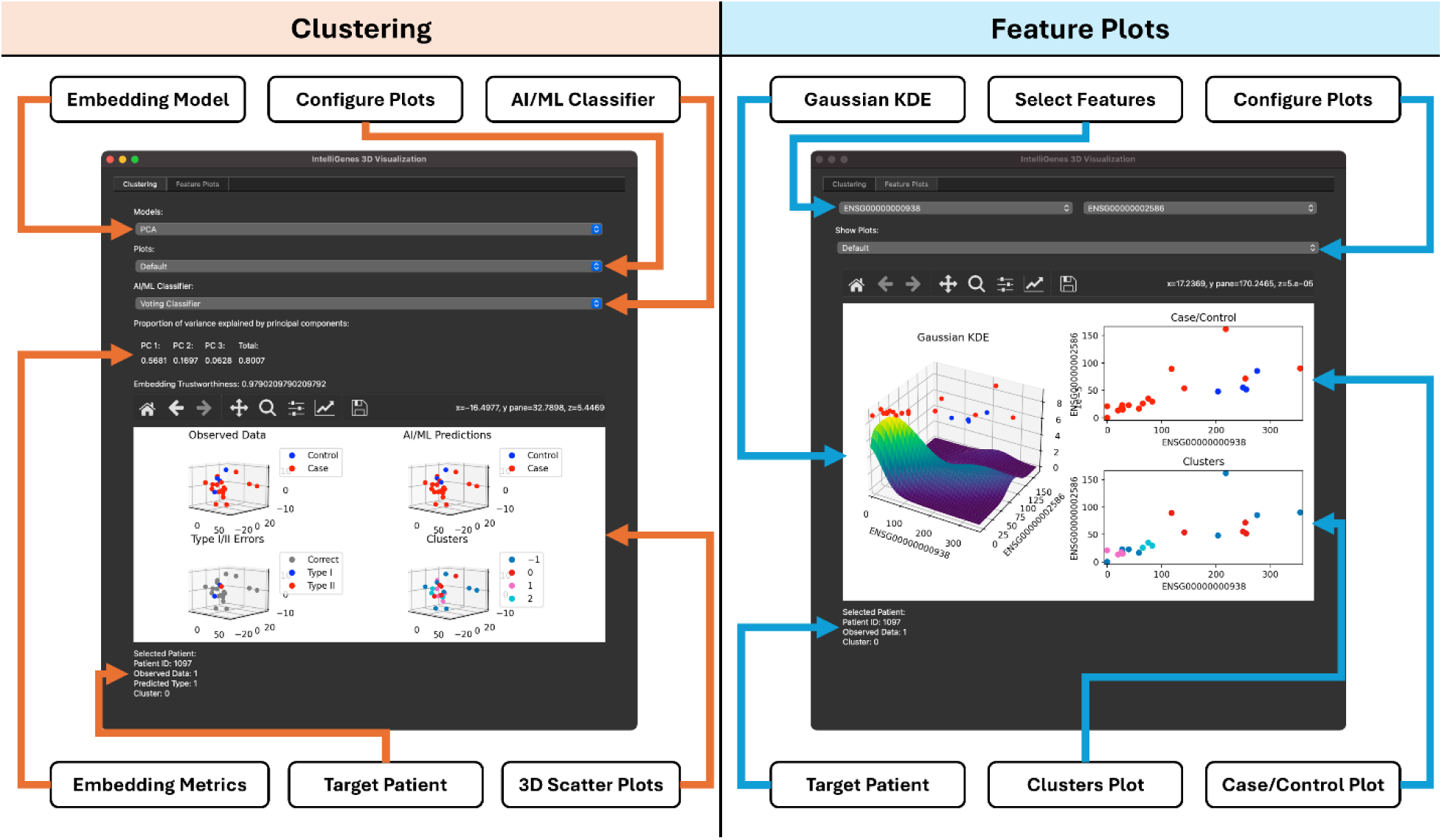
3D *IntelliGenes* interface with Clustering (left) and Feature Plots (right). Clustering enables the configuration of 3D embedding models to view data and AI/ML model performance in 3D. Feature Plots provides density plots and scatter plots to understand inter-feature relationships.

#### Clustering - method

The visualization of high dimensional data in 3D is particularly useful in identifying key structures that may be overshadowed in 2D. Clusters visualized in 3D are superior to their 2D counterparts because the additional dimension often allows for better segregation of classes e.g., indistinguishable diseases states in 2D visualizations may appear more separable when viewed in 3D, may lead to greater understanding of disease-related characteristics. Furthermore, data clustering has the potential to enable the identification of similarities between patients which may be used in downstream investigation to explore correlated pathways. 3D *IntelliGenes* offers data visualization and clustering using a 3D-embedded dataset and enables the user to explore and interact with 3D Visualizations with extensive configuration options. The user is presented with options to select and assess linear and nonlinear embedding models, explore predictions of individual AI/ML classifiers, and analyze patient-specific data. It generates four primary plots: 1) Observed Data, 2) AI/ML Predictions, 3) Type I/II Errors, 4) Clusters. Each of these plots may be independently explored for greater visibility or viewed simultaneously for comparison purposes.

##### 3D Embedding

The visualization of high dimensional data in 3D necessitates embedding the dataset into a 3D vector space, a technique also referred to as dimensionality reduction. To understand the high-level organization of a multi-omics dataset, linear embedding models such as Principal Component Analysis (PCA) are preferred as they preserve global structure by projecting the data into a subspace that minimize the amount of variability lost through dimensionality reduction. However, linear embedding models are ineffective at interpreting highly complex and nonlinear structures in higher dimensions. On the other hand, nonlinear embedding models can better represent these nuanced relationships but offer a skewed view into the underlying global structure. Uniform Manifold Approximation and Projection (UMAP) is a nonlinear embedding model that is effective at preserving local structure [14], making it effective at identifying patient similarities. However, it is unwise to use local structure-preserving models to generalize global patient relationships. Pairwise Controlled Manifold Approximation and Projection (PaCMAP) is a variation of UMAP that aims to strike a middle ground between preserving global and local structure while remaining effective at capturing nonlinear structures [15]. Crucially, each embedding model offers a different view and interpretation into the underlying data organization and structure. In 3D *IntelliGenes*, the user may visualize their CIGT-formatted dataset using any of three different embedding models (i.e., PCA, UMAP, PaCMAP), allowing the user to select the one best tailored to their use case. For the currently selected embedding model, an accuratemetric is reported which indicates the effectiveness of models in preserving the original high-dimensional structure. Moreover, the PCA model also reports how much variability is accounted for through the linear projection.

##### ML Classifiers

To compare the performance and biases of the ensemble of ML models supported by the current *IntelliGenes* methodology, it is crucial to visualize the predictions and errors each classifier makes on a particular dataset. 3D *IntelliGenes* enables the user to explore the individual predictions made by each individual classifier: RF, SVM, XGBoost, KNN, MLP, or the ensembled voting classifier. In doing so, the user can comparatively analyze which ML models perform best on specific regions of data. Within the interface, the user may select a specific classifier or view the default voting classifier. Upon selecting a target classifier, the generated plots automatically update to reflect the predictions and errors of the selected model.

##### Observed Data

The Observed Data plot in 3D *IntelliGenes* shows the distribution of each data point under the currently selected embedding model. It distinguishes between case and control patients, enabling the user to quickly discern how the two classes are related and clustered. Control patients (i.e. patients without the disease state) are labeled in blue, and case patients (i.e. those with the disease state) are labeled in red. 3D *IntelliGenes* also supports the user in inspecting this embedding. Moreover, by hovering over a specific point in any of the scatter matrices, the user is presented with detailed information pertaining to the patient, such as their unique identifier, disease state, predicted disease state using the selected AI/ML classifier, and estimated cluster. The hover preview feature is supported for all clustering plots.

##### AI/ML Predictions

Such visualization supports inspecting the individual predictions made by the currently selected ML classifier. By analyzing such visualization, the user can better understand the accuracy of model as well as assess any biases present in the model. Specifically, regions where a model tends to overpredict a certain disease state could indicate overfitting or the presence of unknown factors that may warrant further investigation. 3D *IntelliGenes* supports the user in visualizing the predictions of each ML classifier, allowing for their comparison within these regions of interest. Consistent with the Observed Data plot, this plot differentiates case and control patients with a unique color: blue points are control patients and red points are case patients. Hovering over individual patients provides similar information about the predicted disease state.

##### Type I/II Errors

Understanding the inaccuracies in the predictions of the ML classifiers is crucial towards assessing and comparing their performance on specific datasets. Although the AI/ML Predictions plot crucially offers a view into the distribution of disease predictions, the specific types of errors made remains obscured. Differentiating between Type I (i.e. false positive) and Type II (i.e. false negative) errors are important towards understanding any inherent imbalance or bias present in each classifier’s predictions. This plot marks correct predictions in gray while highlighting the Type I and Type II errors in blue and red, respectively. Like the 2D confusion matrix heatmap generated by *IntelliGenes GUI*, the user may use this visualization to understand general accuracy and performance of the models. However, unlike a 2D heatmap, this 3D plot allows for simultaneously understanding the structural significance of the misclassifications. Not only can the user inspect what types of errors were made, but also where in the embedded space they occur to determine crucial information, such as whether the misclassification was due to an outlier or because of some other bias.

##### Clusters Plot

To quantify and visualize how patients are biologically similar to each other, 3D *IntelliGenes* supports clustering patients with a density-based algorithm. Identifying points that are more densely packed together might indicate shared biological traits and extracting these clusters may enable the identification and stratification of potential patient populations to explore cluster-specific structures. 3D *IntelliGenes* uses OPTICS, a density-based clustering algorithm which, unlike other clustering algorithms like K-Means or DBSCAN [16], is more resilient to variable density present in the dataset [17]. Effectively clustering patients using density and neighbor-based information can also improve prediction performance on rare alleles [18].

#### Feature plots - method

Interpreting the significance of multi-omics features requires exploring their inter-relationships and visualizing how two features are co-expressed can provide deeper insights into biological pathways underlying their regulation [19]. 3D *IntelliGenes* supports the user in exploring pairwise specific feature plots to visualize the distribution and correlations between biomarkers of interest. In the Feature Plots tab of the interface, the user is presented with options to select two distinct features to visualize. Upon doing so, they are presented with 3 distinct plots combining 3D and 2D into a multi-dimensional approach to support joint feature analysis.

##### Gaussian KDE Plot

Kernel Density Estimation (KDE) is a robust technique used to visualize a smooth probability distribution of samples in a dataset. Unlike a 2D scatter plot where it may be difficult to interpret density and estimate the likelihood of observing new samples in specific regions, a KDE plot more easily distinguishes denser regions in the third dimension through higher peaks and can provide smoother approximations for sparser datasets. 3D *IntelliGenes* generates a KDE plot for a pair of selected features under the assumption of a gaussian (i.e. normal) kernel. In addition, because 3D plots are often more effective when paired with 2D plots to display crucial slices of the 3D structure [20], 3D *IntelliGenes* plots each patient’s disease state (i.e. case/control) in 2D directly above the 3D KDE surface. This enables the user to gain additional information not easily acquirable by separately analyzing each plot, such as simultaneously identifying regions of high-density as well as understanding the distribution of the diseased and healthy patients. When viewed directly from above, this 3D KDE plot has the added effect of appearing as a 2D density heatmap.

##### Case/Control Plot

Although the distribution of disease states is viewable above the 3D KDE plot, 3D *IntelliGenes* also separately generates a flat 2D scatter plot for improved accessibility, navigability. This plot is displayed adjacent to the KDE plot but can be viewed in isolation for better visibility. Consistent with the Clustering module, hovering over each individual point also presents the user with information about the target patient, including their unique identifier, disease state, predicted disease state, and estimated cluster. This hover preview allows for the contextual identification and understanding of each patient.

##### Clusters Plot

For the selected pair of features, 3D *IntelliGenes* also plots the estimated clusters predicted with the density-based OPTICS clustering algorithm as a 2D scatter plot adjacent to the 3D KDE plot. Notably, this plot can be used to assess whether the co-expression of two features is significant in discriminating clusters. In other words, if two distinct clusters have minimal overlap among each other, then the expression of the selected pair of features may help in stratifying the densely packed regions of the original higher dimensional space. Since these clusters may represent some shared biological similarities, such a discovery could indicate a need for further investigation to determine how the biomarker interaction relates to the corresponding biological and disease states.

## Results

Our results are divided into two sections, first is based on the AI/ML-Ready data and algorithms, and other is 3D visualization of results.

### AI/ML-Ready Data and Algorithms

The *IntelliGenes* methodology has been successfully validated in a peer-reviewed study to discover novel biomarkers, predict disease for CVD patients, and create models with high predictive accuracy [2]. Here, we processed a CIGT-formatted dataset with clinical demographics and ubiquitous transcriptomic biomarkers (present in >80% of dataset) from a diverse cohort comprised of 61 CVD patients (40 males and 21 females, aged 45-92) and 10 healthy controls (5 males and 5 females, aged 28-78). All procedures involving human participants were in accordance with the ethical standards of the institution and with the 1964 Helsinki Declaration and its later amendments or comparable ethical standards. All human samples were used in accordance with relevant guidelines and regulations, and all experimental protocols were approved by the Institutional Review Board of Rutgers.

Feature selection was first performed using the default parameters configured in the *IntelliGenes GUI* interface. Namely, Pearson’s correlation, *χ*^2^ test, and ANOVA were all performed features significant at each of the 0.05 thresholds. RFE was performed to extract the top 10% of significant features. Against this criterion, we identified 19 transcriptomic significant biomarkers relevant in predicting the CVD disease state. 5 ML classifiers (i.e. RF, SVM, XGBoost, KNN, MLP) were then trained on 70% of the dataset and tested on the remaining 30%. Independent predictions were then aggregated downstream by a voting classifier. The RF and SVM models performed the best with accuracies of 95% and 91% respectively, and with an area under the ROC curve of around 0.97 each. Combined, the models were able predict CVD disease state in the testing cohort with an accuracy above 86%.

### 3D Visualization

The overall results producing using 3D visualization are based on clustering and feature plots.

#### Clustering - results

##### 3D Embeddings

Dimensionality reduction was performed using 3 embedding models: PCA, UMAP, and PaCMAP (Figure 3). Because dimensionality reduction techniques are highly subject to the variance of individual features, standardization was performed prior to embedding. PCA creates a linear projection onto a 3D space defined by 3 orthogonal principal components. The three principal components explained 57%, 17% and 6% of the variance in the dataset respectively. In other words, 80% of the total variability in the dataset was accounted for through PCA. Moreover, PCA had the highest trustworthiness score of 0.98 indicating that the model preserved most of the pairwise distances of the original dataset. UMAP was performed with the 10 nearest neighbors and a min_dist configuration of 0.1 with lower values resulting in tighter clusters [14]. UMAP had a trustworthiness score of 0.83. Lastly, PaCMAP was configured with the default parameters of mid-near ratio of 0.5 and a further pair ratio of 2 [15]. PaCMAP had a trustworthiness score of 0.85. Expectedly, PCA had the most trustworthy embedding as linear projections tend to aim global structure. UMAP had the least trustworthy embedding as it is designed to preserve local pairwise distance over global structure.

**Figure 3.**
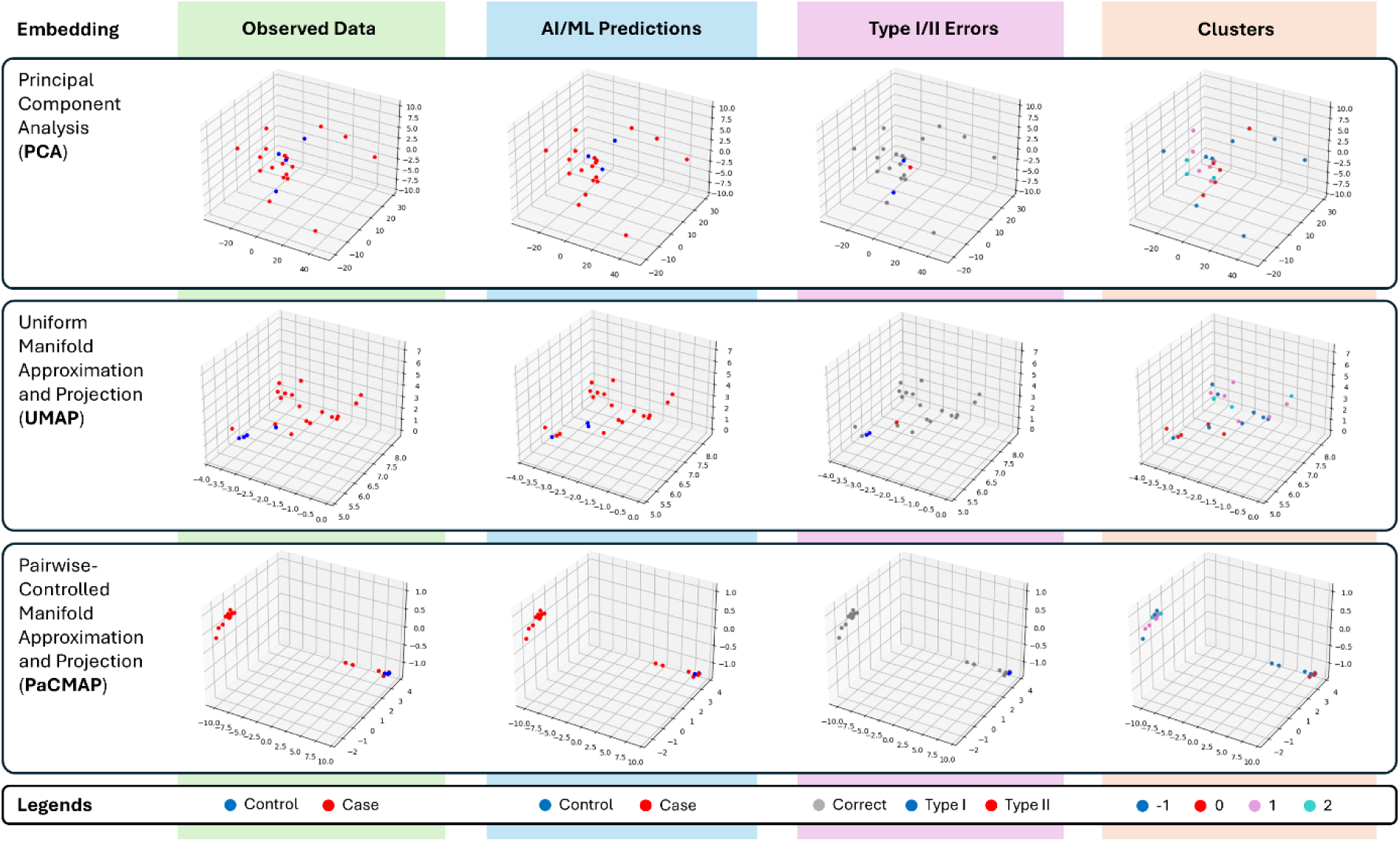
Clustering results on CVD cohort. Organizes 3D scatter matrices representing Observed Data, AI/ML Predictions, Type I/II Errors, and Clusters, for 3 separate embeddings (i.e. PCA, UMAP, PaCMAP). Each point represents a different patient in the test cohort.

##### ML Classifiers

Five separate ML classifiers (i.e. RF, SVM, XGBoost, KNN, MLP) were trained using 70% of the cohort, and validated on the remaining 30%. Individual predictions were ensembled by a voting classifier by considering individual classifier prediction probabilities. The ensembled voting classifier performed with an accuracy of around 86%, F1 score of 0.86 and an area under the ROC curve (ROC-AUC) of 0.94. Individually, we see that RF and SVM performed the best with 95% and 91% accuracy, and F1 scores of 0.96 and 0.91, respectively. Comparatively, the MLP classifier performed worst on the dataset, achieving an ROC-AUC of 0.64. The remaining classifiers all achieved ROC-AUC scores above 0.88 demonstrating high discriminatory capabilities.

##### Observed Data Plot

This plot visualizes the 18 case patients and 4 control patients in our testing cohort under each of the 3 embedding models, highlights their differences in discriminating between disease states. Qualitatively, we see that the nonlinear embedding models (i.e. PaCMAP, UMAP) are better able to split the dataset into separate clusters based on case/control status, with PaCMAP best able to create visually discernible clusters with minimal overlap. Analyzing the scatter distribution produced by each dimensionality reduction technique provides a more holistic understanding of the underlying dataset with healthy patients being segmented from patients with CVDs.

##### Predicted Data Plot

To better understand the performance of each ML classifier as well as the ensembled voting classifier, it is imperative to visually compare the distribution of the predicted disease states. Through this plot, one can gain a general intuition for the decision boundary for each classifier as well as the comparative bias of each model. For example, we see that tree-based models like RF and XGBoost seem to predict generally the same disease state within similar embedding regions. On the other hand, the KNN classifier was more volatile, with predicted healthy patients having greater variance under the PCA embedding. Generally, we also see that the ensembled voting classifier more accurately partition the 3D space into regions of healthy patients with less overall variance than each individual model.

Type I/II Errors Plot: Analyzing the errors made by each model is crucial towards understanding inherent bias and limitations. A Type I error is a false positive (i.e. predicted the disease state when the patient was healthy) and a Type II error is a false negative (i.e. predicted healthy when the patient is diseased). Interestingly, we see that the tree-based models (e.g. RF, XGBoost) only made Type I errors on our dataset, with RF making a single mistake and XGBoost making 4 misclassifications. The other models seem to predict roughly equal Type I and Type II errors, with SVM making one each, and KNN and MLP making two each. Additionally, we see that the errors made by each model tend to overlap, for example both RF and SVM misclassify the same patient.

##### Clusters Plot

Clustering is performed using a density-based OPTICS model, configured to include at least 3 samples in each cluster. On our dataset, OPTICS identifies 3 clusters numerically labeled 0, 1, and 2. The −1 label is used to denote samples that do not belong to any specific cluster typically due to being outliers or spaced evenly between other clusters. Notably, each of the embedding models generally preserves the high-dimensional clustering, with cluster 0 (colored red) being more dissimilar compared to cluster 1 (colored pink) and cluster 2 (color cyan). Further investigation is required to understand any biological phenomenon particularly relevant within each cluster. Additionally, further differential analysis is required to segregate the patients labeled as −1 by OPTICS.

#### Feature plots - results

In the study employing *IntelliGenes* to predict CVD in a diverse cohort, we identified four primary features that were significant across multiple classifiers including 3 biomarkers with previously studied disease associations (i.e., *MTRNR2L1*, *HLA-B*, *LILRA2*), and one novel biomarker (i.e., *RN7SL593P*) [2]. Specifically, *MTRNR2L1* was the best predictor across models, being ranked in the top three across SVM, XGBoost, and KNN classifiers. *HLA-B* has been previously implicated in cardiomyopathy [21], *LILRA2* has been associated with coronary artery disease [22], and *MTRNR2L1* has been found to be associated with patients with breast cancer, another prevalent condition of patients in our original CVD dataset. No direct links to disease have been studied for *RN7SL593P*, warranting further investigation into its role in CVD manifestation. To perform joint feature analysis using 3D *IntelliGenes*, we analyzed the inter-relationships of these 4 genes (Figure 4). Since *MTRNR2L1* was the most heavily relied upon biomarker during prediction, we analyzed its joint expression with the novel biomarker *RN7SL593P*. Similarly, we investigated the relationships between *LILRA2* and *HLA-B*, as both were biomarkers with previously studied links to CVDs. Results presented in our dataset are encoded using Ensembl IDs [23], instead of gene symbols. The corresponding IDs for the four relevant biomarkers are: *MTRNR2L1* is ENSG00000256618, *RN7SL593P* is ENSG00000266422, *HLA-B* is ENSG00000234745, and *LILRA2* is ENSG00000239998. A similar analysis should be conducted between all pairs of interesting genes to gain a deeper insight into more global interactions.

**Figure 4.**
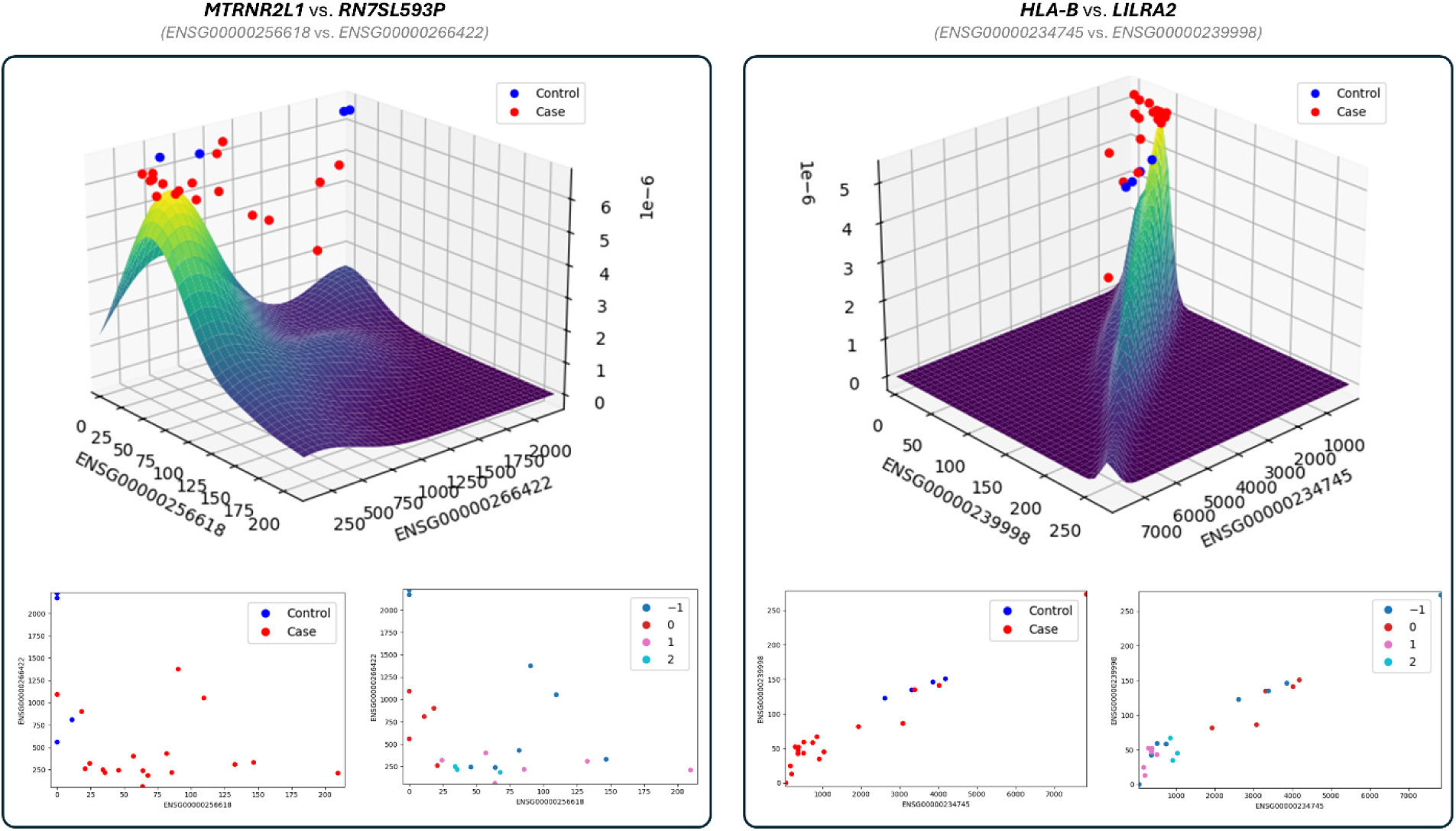
Pairwise Feature Plots results, including density plots and scatter plots, for select pairs of features: *MTRNR2L1* vs. *RN7SL593P* (left) and *HLA-B* vs. *LILRA2* (right). Each point represents a different patient in the test cohort, and the x and y axes represent the two features.

##### Gaussian KDE Plot

The KDE surface for each pair of investigated genes helps highlighting the interesting relationships. We see that the joint surface of *MTRNR2L1* and *RN7SL593P* is unimodal indicating that the co-expression of both genes in the dataset appears roughly normally distributed centered. The single mode is centered around an expression value of approximately 50 TPM for *MTRNR2L1* and approximately 300 TPM for *RN7SL593P*. The joint distribution of *HLA-B* and *LILRA2* within our dataset is also particularly interesting. Here, we see a stark linear correlation between the two genes, expressed by a narrow ridge in the estimated surface. Notably, both CVD associated genes seem to have strong co-expression, warranting further investigation into their shared functional pathways.

##### Case/Control Plot

Although the unimodal distribution between *MTRNR2L1* and *RN7SL593P* is less evident in the 2D scatter plot, this plot highlights additional information about the distribution of the case and control patients themselves. Particularly, we see that the upregulation of *MTRNR2L1* and the downregulation of *RN7SL593P* seems to be more correlated with the CVD disease state. In other words, the healthy patients appear to minimally express *MTRNR2L1* and more substantially express *RN7SL593P*. On the other hand, the co-expression of the CVD-associated genes (i.e., *HLA-B* and *LILRA2*) have inseparable classes, likely indicating the role of other biomarkers in discriminating the disease states. Nonetheless, we observe that the healthy patients in our dataset are centrally clustered with the case patients appearing with both downregulated and upregulated genes.

##### Clusters Plot

This clusters plot is beneficial in understanding how similarly identified patients are distributed across the two pairs of investigated genes. Notably, between *HLA-B* and *LILRA2*, the identified clusters appear generally separable demonstrating the potential of these CVD-associated genes in stratifying the patient population. Cluster 0 (red) appears to have an upregulated expression for both genes, while clusters 1 (pink) and 2 (cyan) both have generally downregulated expressions corresponding to diseased patients. The cluster plot between *RN7SL593P* and *MTRNR2L1* is less interesting with high variability among the identified clusters. Moreover, clusters 1 and 2 have high overlap indicating that the co-expression of these two genes is less relevant in understanding the global structure of the dataset.

## Discussion

The dynamic and interactive visualization of multi-omics data in 3D has the potential to augment data interpretability, render it more accessible to the wider scientific community, and provide a deeper understanding of high-level trends and relationships [6]. With the application of various AI/ML techniques to omics data, it is imperative to develop novel tools to enhance the analytical skills of the research workforce. There is a notable lack of robust and reliable visualization tools to aid in the interpretation and understanding of these black-box results generated by modern AI/ML analysis. Portable, interoperable, and user-friendly software is required to drive community engagement in using, testing, replicating, and validating scientific results.

Current state-of-the-art tools for 3D visualizations employ dimensionality reduction and clustering to identify patterns but are limited in understanding feature-specific trends for multi-omics and AI/ML related metrics [6, 24]. Here, we developed and proposed 3D *IntelliGenes*, an extension of our *IntelliGenes* methodology offering interactive 3D visualization options to enhance the data exploration and interpretation workflow. 3D *IntelliGenes* was developed to promote FAIR4RS principles by supporting scientists of varying bioinformatic backgrounds in interacting with and visualizing integrated multi-omics data.

In the future, 3D *IntelliGenes* may be extended to offer a more diverse suite of 3D visualizations for better understanding the nuances of multi-omics data. 3D graphics are not only impactful when combined with more widely accessible 2D plots [20] but also have potential for a much greater set of unique interactive visualization options. Advancements in AR/VR technologies offer an immersive and physically interactive 3D digital space through which data can be better explored and understood [25]. Although it remains widely unexplored for data visualization, AR/VR holds great potential in transforming visualization in medical science, and further research in this field can elucidate greater understanding of diseases and their treatments [25]. Tools like 3D *IntelliGenes* have the potential to disseminate complex AI/ML data analysis and visualization to a greater scientific audience and can aid in improving the analytical and interpretation skills of the broader scientific community.

## Conclusions

In the era of precision medicine, the generation, analysis, and visualization of multi-omics data holds great promise for uncovering complex biological pathways. The visualization of integrated multi-omics data in 3D can streamline the exploration, analysis, evaluation, and interpretation of AI/ML results, yet remains critically underexplored in literature. The development of findable, accessible, interoperable, and reproducible software tools like 3D *IntelliGenes* has potential to impact researchers and physicians in clinical settings by aiding in the identification of novel and predictive biomarkers to enable earlier diagnosis and personalized treatment options for complex diseases. The presented results in this study include 3D scatter matrices demonstrating disease state distribution, case/control predictions of various AI/ML models, type I/II error tendencies, and density-driven patient clustering, as well as two joint analyses of four carefully selected biomarkers with high predictive power as published in a recent peer-review study of ours (i.e., *MTRNR2L1*, *HLA-B*, *LILRA2*, *RN7SL593P*).

## Data Availability

The source code of 3D IntelliGenes is publicly available at GitHub.

https://github.com/drzeeshanahmed/3D-IntelliGenes

## Abbreviations

ANOVA: Analysis of Variance
AI: Artificial Intelligence
AR: Augmented Reality
CVD: Cardiovascular Disease
*χ*^2^: Chi-Squared
CIGT: Clinically Integrated Genomics and Transcriptomics
FAIR4RS: Findable, Accessible, Interoperable, Reproducible for Research Software
GUI: Graphical User Interface
I-Gene: Intelligent Gene
KNN: K-Nearest Neighbors
KDE: Kernel Density Estimation
ML: Machine Learning
MLP: Multi-Layer Perceptron
PaCMAP: Pairwise-Controlled Manifold Approximation and Projection
PCA: Principal Component Analysis
RF: Random Forest
ROC: Receiver Operating Characteristic curves
RFE: Recursive Feature Elimination
ROC-AUC: ROC Area Under Curve
SHAP: Shapley Additive exPlanations
SVM: Support Vector Machine
3D: Three-Dimensional
2D: Two-Dimensional
UMAP: Uniform Manifold Approximation and Projection
VR: Virtual Reality
WGS: Whole Genome Sequencing
XGBoost: Xtreme Gradient Boosting

## Declarations

### Ethical Approval and Consent to participate

Informed consent was obtained from all subjects. All human samples were used in accordance with relevant guidelines and regulations, and all experimental protocols were approved by the Institutional Review Board.

### Consent for publication

Not applicable

### Availability of data and material

The source code of 3D *IntelliGenes* is publicly available at GitHub <https://github.com/drzeeshanahmed/3D-IntelliGenes>

## Acknowledgments

We appreciate great support by the Department of Medicine, Robert Wood Johnson Medical School; Rutgers Institute for Health, Health Care Policy, and Aging Research; and Rutgers Health, at Rutgers, The State University of New Jersey. We thank members and collaborators of Ahmed Lab for their support, participation, and contribution to this study.

## Author contributions

ZA proposed, supported, allocated resources, and led the *IntelliGenes* project and overall study as the Principal Investigator (PI) and Project Director (PD). RN and WD participated in the AI/ML design and development activities. EP supported post-development activities including overall administration and evaluation of results. SZ provided collaborated support to the overall project. All authors participated in writing of the manuscript and have reviewed and approved it for the publication.

## Biographical Note

RN, EP, and WD are the Research Assistant at the Ahmed lab, Rutgers IFH/RWJMS.

DM is the Software Engineer at the Ahmed lab, and Computer and Information Research Scientist: Programmer Analyst I at the Rutgers IFH.

SZ is the Research Assistant Professor at the Department of Biomedical and Health Informatics, at UMKC School of Medicine.

ZA is the Assistant Professor at the Department of Medicine, Division of Cardiovascular diseases and Hypertension at Rutgers RWJMS. ZA is a Core Faculty Member at the Rutgers Institute for Health.

